# Association between Anemia and Hematologic Traits at Hospital Discharge with One-Year Mortality Among Survivors of Critical Illness

**DOI:** 10.64898/2026.08.03.26359579

**Authors:** Lucas L. Marinho, Danielle Jeong, Maya D’Angelo, John C. Marshall, James A. Russell, Zoe McQuilten, Alisa M. Higgins, Dennis T. Ko, Olivia Haldenby, Jiming Fang, Adriana Luk, Maira Souza-Silva, Marie C. Ferland, Gloria Mutombo, Jordyn Nadler, Felipe Melo Nogueira, Remo Holanda, Sean van Diepen, Ryan Zarychanski, Johnathan P. Mack, Patrick R. Lawler

## Abstract

**Background:** Molecular markers and mediators of adverse post-hospital outcomes following critical illness remain incompletely characterized. Anemia and red blood cell (RBC) indices integrate inflammation, nutritional status, and hematopoietic function, and may represent biological processes that influence long-term outcomes. We therefore examined whether pre-discharge anemia and hematologic clusters derived from correlated RBC indices were associated with one-year mortality among critical illness survivors.

**Methods:** We conducted an exploratory, retrospective cohort study of adult ICU patients discharged alive using MIMIC-IV. Pre-hospital discharge RBC indices (hemoglobin, red cell distribution width (RDW), mean corpuscular volume (MCV), mean cell hemoglobin (MCH), mean cell hemoglobin concentration (MCHC), and RBC count) were analyzed. Hemoglobin and anemia were first evaluated, followed by Gaussian mixture modeling of standardized RBC indices to identify patient clusters in a derivation cohort (70%) and validated findings in a held-out cohort (30%). Associations with one-year mortality were assessed using multivariable-adjusted Cox models. Results:

Among 20,233 ICU survivors with available pre-discharge hemoglobin (median age, 67 years), based on the WHO definition of anemia, 17,247 (85.2%) critical illness survivors were anemic at hospital discharge. Anemia was associated with higher one-year mortality (adjusted HR 1.42, p<0.001). Mortality was 20% overall and decreased across increasing hemoglobin quartiles, from 28.8% to 12.5% (log-rank P<0.005); lower hemoglobin was associated with progressively higher adjusted one-year mortality risk. Six hematologic clusters derived from RBC indices were identified with one-year mortality ranging from 10.1% to 31.4% (log-rank p<0.005). Compared with the lowest-risk group, the cluster characterized by elevated RDW and MCV had a twofold higher adjusted mortality risk (HR 2.14; 95% CI, 1.81–2.53).

**Conclusion:** Both anemia and hematologic clusters derived from standard RBC indices at hospital discharge are associated with one-year mortality following critical illness. These hypothesis-generating findings support further investigation of RBC indices as potential biomarkers for post-discharge risk stratification and of the underlying biological pathways as possible therapeutic targets.

## Background

Long-term outcomes among intensive care unit (ICU) survivors remain poor, with many survivors experiencing rehospitalization or death within one year.[1–4] Few prognostic biomarkers are available to stratify long-term risk at hospital discharge. Their identification may improve post-discharge mortality risk assessment, indicate mediators of later mortality, and identify novel therapeutic targets.

Red blood cell (RBC) indices reflect age, sex, comorbidities, nutritional status, inflammation, and overall marrow function.[5–7] Derangements in these biological processes are common to multiple critical illness syndromes. However, these parameters at hospital discharge following critical illness, when RBC indices may reflect chronic disease, in-hospital treatment, residual biological stress, and/or impaired hematopoietic recovery, have received scant attention. RBC indices at discharge may capture potentially treatable traits (e.g., anemia) and guide tailored interventions in ICU survivors. Such an approach has improved long-term outcomes among patients with chronic heart failure and chronic kidney disease.[8, 9]

Because RBC indices are intercorrelated measures with overlapping distributions, individual parameters do not fully capture the complexity of erythroid physiology. Hematologic clusters, defined by patterns across routinely measured complete blood count (CBC)-derived RBC indices, may identify patient groups with differing erythroid physiology.[10] Such clusters have not been characterized at hospital discharge, and their relationship to long-term adverse outcomes has not been studied. Data-driven clustering of pre-discharge RBC indices may reveal biologically meaningful clusters and provide insight into persistent physiological disturbances associated with long-term risk.[10]

We hypothesized that anemia as well as hematologic clusters derived from correlated RBC indices measured at hospital discharge would be associated with one-year mortality for ICU survivors. In this exploratory analysis, we evaluated these hypotheses using routinely-measured pre-discharge hemoglobin, and also performed data-driven patient clustering based on pre-discharge routine RBC indices as part of the CBC in a large single-center cohort of adult ICU survivors.

## Methods

### Data Source

Data were obtained from the Medical Information Mart for Intensive Care IV (MIMIC-IV) database, version 3.1, a publicly available de-identified electronic health record database of all ICU admissions at Beth Israel Deaconess Medical Center (Boston, USA), from 2008 to 2022.[11] The database includes demographics, laboratory results, vital signs, ICD-9 and ICD-10 diagnoses, procedures, and outcomes, encompassing 94,458 ICU admissions. MIMIC-IV was approved by the Massachusetts Institute of Technology and BIDMC institutional review boards with a waiver of informed consent. This study was further approved by the McGill University Health Centre Research Ethics Board (2026-12503).

### Study Population

We identified all adult (≥18 years) ICU patients discharged alive during the study period, and included only the index ICU admission for those with multiple hospitalizations or ICU readmissions. To ensure meaningful exposure to critical illness, we restricted the cohort to patients with an ICU length of stay ≥ 48 hours and excluded elective or same-day surgical admissions. We further excluded patients who were discharged against medical advice, were transferred to another acute care facility, or lacked complete discharge information. Patients with all RBC pre-discharge parameters missing were excluded.

### Baseline and Critical Illness Characteristics

Baseline characteristics were obtained within 24 hours of ICU admission. Comorbidities were identified using validated ICD codes.[12] Acute critical illness severity was characterized using the highest Sequential Organ Failure Assessment (SOFA) and Oxford Acute Severity of Illness Score (OASIS) values within 24 hours of ICU admission. Sepsis at ICU admission was defined according to Sepsis-3 criteria, using validated MIMIC-IV algorithms.[13] Surgical procedures during hospitalization were identified using the Agency for Healthcare Research and Quality procedure classification system, which maps ICD-9 and ICD-10 procedure codes into surgical and non-surgical. RBC transfusions were recorded throughout the hospitalization. Indicators of organ support were used to characterize critical illness severity and included invasive mechanical ventilation and any vasopressor use in the first 48 hours.

### Exposures: Pre-discharge RBC Measures

Pre-discharge hematologic parameters were assessed using CBC measurements obtained within 72 hours prior to hospital discharge. When multiple measurements were available, the value closest to discharge was selected.

The primary exposure of interest was anemia assessed using pre-discharge hemoglobin levels. Anemia was defined based on the World Health Organization (WHO) sex-stratified cut-offs (<12.0 g/dL for nonpregnant women and <13.0 g/dL for men).[14]

The secondary exposure was hematologic clusters derived from correlated RBC indices, including hemoglobin, red cell distribution width (RDW), mean corpuscular volume (MCV), mean cell hemoglobin (MCH), mean cell hemoglobin concentration (MCHC), and RBC count. These indices capture complementary dimensions of erythrocyte biology: MCV reflects RBC size, MCH hemoglobin content per red blood cell, MCHC hemoglobin concentration within RBC, and RDW variability in RBC size. One patient with missing pre-discharge hemoglobin data was excluded from the primary exposure analysis. Missingness for other pre-discharge RBC indices was also infrequent (<1%). For the clustering analysis, missing RBC values were imputed using chained equations with Bayesian ridge regression and posterior sampling, generating a single completed dataset that was subsequently used for the clustering analysis. Imputed values were used solely for clustering procedures; outcome and covariate data were not imputed.

### Outcomes

The primary outcome for association studies was all-cause post-hospital discharge mortality within one year. Mortality status was ascertained in MIMIC-IV using linked hospital records and state vital statistics.[11] Although this approach is widely used, mortality ascertainment relies on state rather than national records; therefore, some out-of-state deaths may not have been captured. This underascertainment may have been nonrandom, potentially introducing differential outcome misclassification.

### Statistical Analysis

#### Pre-Hospital Discharge Anemia

We examined associations between pre-discharge hemoglobin and 1-year mortality using multivariable Cox regression models. Kaplan–Meier curves were constructed to visualize estimated cumulative survival across pre-discharge hemoglobin quartiles, with differences assessed by log-rank tests. Hemoglobin was also modeled continuously using restricted cubic splines with five knots,[15] using the median as the reference. The five knots were selected to balance flexibility and parsimony in modeling non-linear effects and were placed at the recommended percentiles (5th, 27.5th, 50th, 72.5th, and 95th).[16]

Multivariable Cox proportional hazards models were used to estimate hazard ratios (HR) with 95% confidence intervals (CI) for one-year mortality associated with pre-discharge hemoglobin, adjusting for prespecified variables associated with post-discharge survival, including: demographic (age, sex, weight); baseline clinical and laboratory tests (Charlson comorbidity index, severe liver disease, cancer, sepsis diagnosis, surgical procedure, maximum creatinine value in the first 24hr); and critical illness severity markers during hospitalization (ICU and hospital length of stay, SOFA, Oasis, renal replacement therapy, vasopressor use in the first 48hr, RBC transfusion volume). The proportional hazards assumption was assessed using Schoenfeld residual tests.

#### Erythroid Clustering Approach (Derivation Cohort)

For the clustering analysis, the cohort was randomly split 70:30 into derivation and internal hold-out validation cohorts. Clustering was performed in the derivation cohort, whereas the validation cohort was used to assess clusters reproducibility and outcome associations.

We used a probabilistic, model-based clustering approach to identify hematologic clusters derived from correlated pre-discharge RBC indices. Gaussian mixture models (GMMs) were fitted using standardized (z-score–transformed) RBC variables. Because RBC indices are continuous, intercorrelated measures with overlapping distributions, GMMs were selected as they permit flexible covariance modeling and probabilistic assignment that may better reflect biological heterogeneity. Models with two to six clusters were compared, and the optimal number of clusters was selected based on the lowest Bayesian Information Criterion. Individual patients were assigned to clusters according to maximum posterior probability. To assess the stability of cluster number selection, a bootstrap resampling was used, while pairwise adjusted Rand indices evaluated clustering reproducibility.

#### Internal Validation

Patients in the internal held-out validation cohort were assigned to clusters using posterior probabilities derived from the final GMM fitted in the derivation cohort. Internal cluster validity and separation were assessed using the Silhouette coefficient and Davies–Bouldin index.

Associations between cluster membership and outcomes were evaluated separately in the derivation and validation cohorts using identical statistical models and covariate adjustment sets.

Cluster Characterization and Analysis

Baseline characteristics were compared across clusters using analysis of variance or Kruskal– Wallis tests for continuous variables, as appropriate, and χ² tests for categorical variables.

Cluster interpretation was informed by patterns of RBC indices and associated clinical characteristics.

Kaplan-Meier curves, log-rank tests, adjusted Cox models, and Schoenfeld residual tests were similarly applied here to compare 1-year survival across erythroid clusters derived from correlated RBC indices in the derivation and validation cohorts.

#### Software and Codes

Analyses were performed using Python (v3.10) with pandas, NumPy, scikit-learn, statsmodels, SciPy, lifelines, matplotlib, seaborn, and umap-learn.

## Results

### Patients and Analysis Cohorts

Of 94,458 ICU admissions from the MIMIC-IV database, 20,234 adults were eligible for the study (Figure 1). The median age was 67 years (IQR 55-78), and 55.9% were male (Table 1). Diabetes mellitus was the most common comorbidity (33.8%). The median maximum SOFA score within the first 24 hours was 4 (IQR, 2–7), and the median hospital length of stay was 10 days (IQR, 6– 16) (Table 1). Overall, 44.1% of patients met Sepsis-3 criteria at admission, 52.4% underwent a surgical procedure during hospitalization, and 57.2% required invasive mechanical ventilation (Table 1). For cluster analysis, patient characteristics were similar between the derivation (n= 14,163) and validation (n= 6,071) groups (eTable 1).

**Figure 1.**
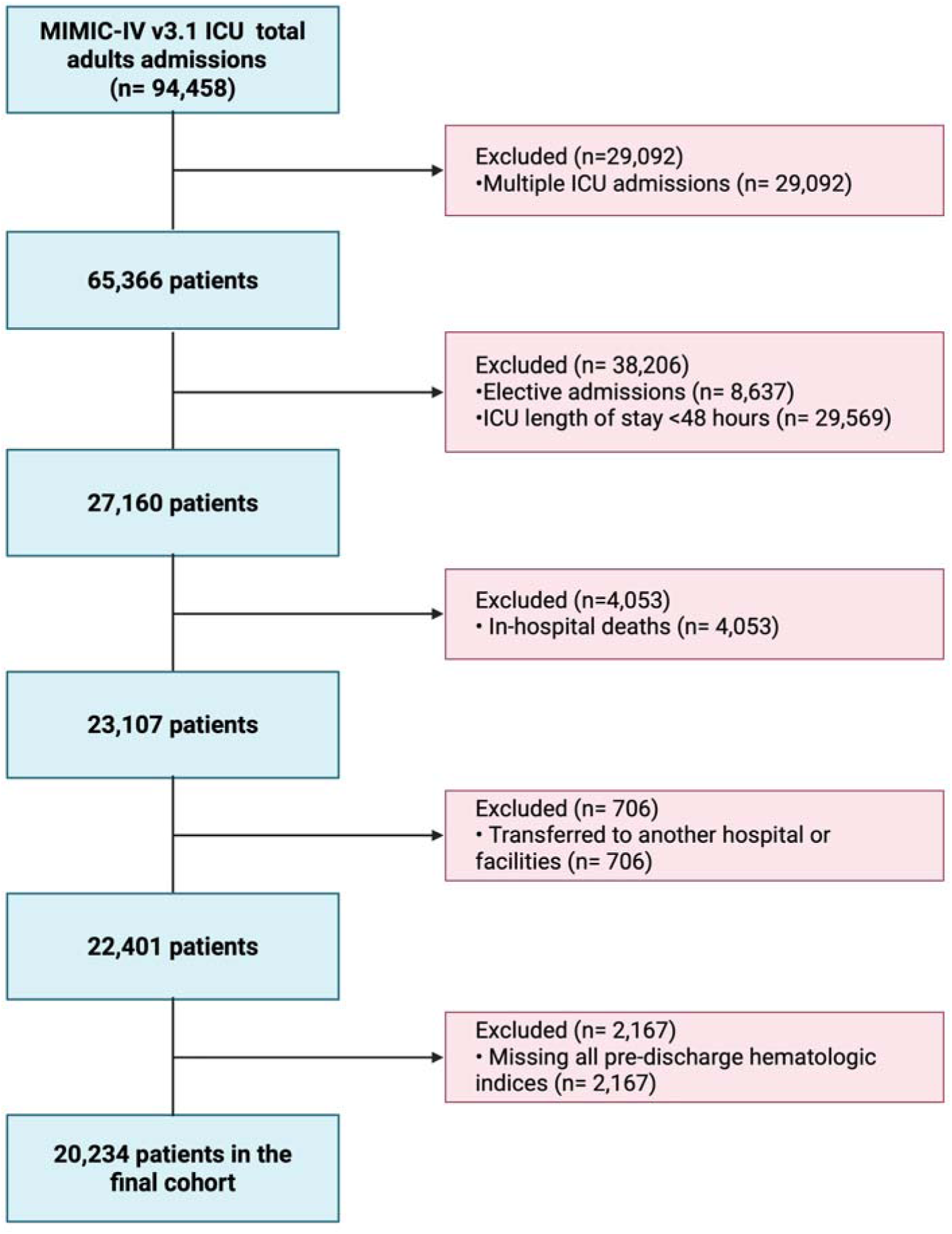
**Cohort creation**

**Table 1.**
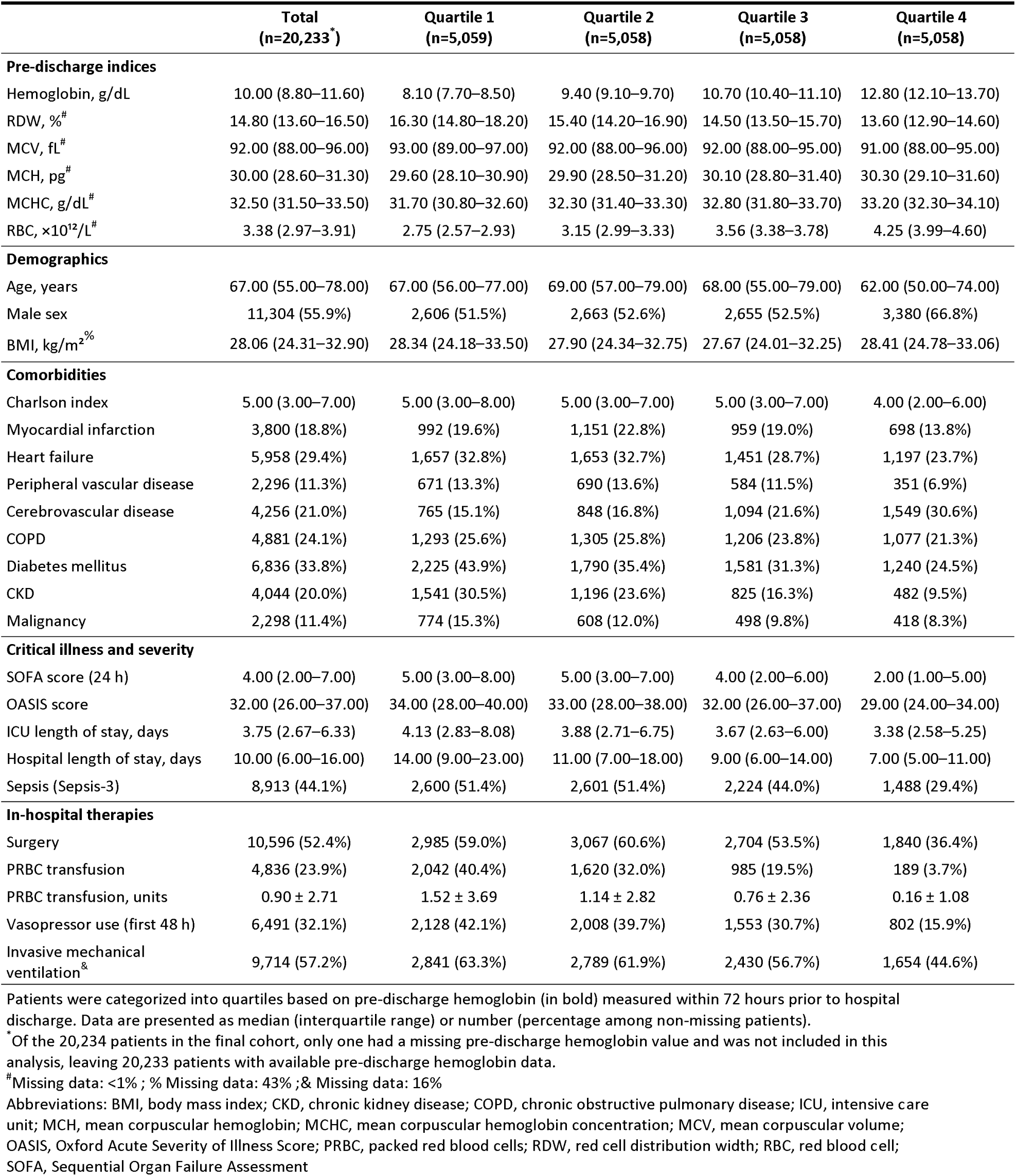
Clinical characteristics of critical illness survivors across quartiles of pre-discharge hemoglobin.

**Table 1.** Baseline characteristics by hematologic clusters derived from RBC indices at discharge in the derivation cohort.

| Characteristic | Cluster 0 (n=3,607) | Cluster 1 (n=1,087) | Cluster 2 (n=2,656) | Cluster 3 (n=929) | Cluster 4 (n=3,088) | Cluster 5 (n=2,796) |
| --- | --- | --- | --- | --- | --- | --- |
| **Pre-discharge indices** |  |  |  |  |  |  |
| Hemoglobin, g/dL | 8.60 (8.10–9.20) | 9.10 (8.10–10.70) | 12.80 (12.00–13.80) | 10.30 (8.90–12.10) | 10.90 (10.40–11.70) | 9.40 (8.70–10.00) |
| RDW, % | 15.70 (14.40–17.30) | 16.30 (14.20–19.15) | 13.60 (13.00–14.40) | 17.20 (15.30–19.90) | 14.00 (13.20–14.90) | 15.60 (14.50–16.90) |
| MCV, fL | 95.00 (92.00–97.00) | 103.00 (100.00–107.0) | 90.00 (87.00–92.00) | 80.00 (77.00–84.00) | 94.00 (91.00–97.00) | 88.00 (85.00–91.00) |
| MCH, pg | 30.40 (29.60–31.20) | 33.70 (32.80–34.80) | 29.70 (28.80–30.60) | 25.00 (23.40–26.20) | 31.10 (30.30–31.90) | 28.10 (27.20–28.80) |
| MCHC, g/dL | 32.10 (31.30–33.10) | 32.90 (31.80–33.90) | 33.10 (32.30–33.90) | 31.10 (29.90–32.00) | 33.20 (32.40–34.00) | 31.80 (30.90–32.70) |
| RBC, $\times 10^{12}/L$ | 2.85 (2.69–3.01) | 2.70 (2.43–3.13) | 4.30 (4.10–4.58) | 4.13 (3.73–4.81) | 3.54 (3.35–3.74) | 3.32 (3.13–3.54) |
| **Demographics** |  |  |  |  |  |  |
| Age, years | 69.00 (58.00–79.00) | 64.00 (54.00–76.00) | 62.00 (48.00–74.00) | 63.00 (50.00–75.00) | 68.00 (55.00–79.00) | 68.00 (56.00–78.00) |
| Male sex | 1,937 (53.7%) | 632 (58.1%) | 1,736 (65.4%) | 489 (52.6%) | 1,731 (56.1%) | 1,372 (49.1%) |
| BMI, kg/m<sup>2</sup> | 27.79 (24.01–32.69) | 27.10 (23.28–31.33) | 28.63 (25.00–33.39) | 30.35 (25.75–38.09) | 27.39 (23.94–31.83) | 28.60 (24.72–33.95) |
| **Comorbidities** |  |  |  |  |  |  |
| Charlson index | 5.00 (3.00–7.00) | 5.00 (3.00–7.00) | 4.00 (2.00–6.00) | 5.00 (2.00–7.00) | 4.00 (2.00–6.00) | 5.00 (3.00–7.00) |
| Myocardial infarction | 790 (21.9%) | 150 (13.8%) | 350 (13.2%) | 115 (12.4%) | 590 (19.1%) | 610 (21.8%) |
| Heart failure | 1,163 (32.2%) | 298 (27.4%) | 572 (21.5%) | 362 (39.0%) | 793 (25.7%) | 976 (34.9%) |
| Peripheral vascular disease | 526 (14.6%) | 96 (8.8%) | 160 (6.0%) | 80 (8.6%) | 338 (10.9%) | 350 (12.5%) |
| Cerebrovascular disease | 559 (15.5%) | 182 (16.7%) | 835 (31.4%) | 182 (19.6%) | 705 (22.8%) | 506 (18.1%) |
| COPD | 891 (24.7%) | 277 (25.5%) | 518 (19.5%) | 292 (31.4%) | 667 (21.6%) | 761 (27.2%) |
| Diabetes mellitus | 899 (24.9%) | 193 (17.8%) | 554 (20.9%) | 309 (33.3%) | 616 (19.9%) | 807 (28.9%) |
| CKD | 1,059 (29.4%) | 233 (21.4%) | 237 (8.9%) | 179 (19.3%) | 411 (13.3%) | 690 (24.7%) |
| Malignancy | 502 (13.9%) | 171 (15.7%) | 220 (8.3%) | 117 (12.6%) | 286 (9.3%) | 351 (12.6%) |
| **Critical illness and severity** |  |  |  |  |  |  |
| SOFA score (24 h) | 5.00 (3.00–8.00) | 6.00 (3.00–9.00) | 2.00 (1.00–4.00) | 3.00 (2.00–5.00) | 4.00 (2.00–6.00) | 4.00 (2.00–7.00) |
| OASIS score | 34.00 (28.0–39.0) | 33.00 (27.0–38.0) | 28.00 (23.0–34.0) | 30.00 (25.0–36.0) | 31.00 (26.0–37.0) | 33.00 (27.0–38.0) |
| ICU LOS, days | 4.04 (2.75–7.75) | 3.96 (2.83–6.96) | 3.33 (2.54–5.25) | 3.58 (2.63–5.79) | 3.54 (2.63–5.92) | 3.92 (2.75–6.68) |
| Hospital LOS, days | 13.00 (8.00–22.0) | 12.00 (7.00–20.0) | 7.00 (5.00–11.00) | 9.00 (6.00–15.00) | 9.00 (6.00–13.00) | 11.00 (7.00–18.0) |
| Sepsis (Sepsis-3) | 1,860 (51.6%) | 566 (52.1%) | 775 (29.2%) | 354 (38.1%) | 1,311 (42.5%) | 1,367 (48.9%) |
| **In hospital therapies** |  |  |  |  |  |  |
| Surgery | 2,295 (63.6%) | 459 (42.2%) | 1,009 (38.0%) | 338 (36.4%) | 1,674 (54.2%) | 1,649 (59.0%) |
| PRBC transfusion | 1,451 (40.2%) | 283 (26.0%) | 123 (4.6%) | 139 (15.0%) | 564 (18.3%) | 809 (28.9%) |
| PRBC transfusion, units | 1.64 ± 3.79 | 1.06 ± 3.40 | 0.19 ± 1.10 | 0.36 ± 1.23 | 0.71 ± 2.11 | 0.93 ± 2.26 |
| Vasopressor use (first 48 h) | 1,553 (43.1%) | 353 (32.5%) | 420 (15.8%) | 215 (23.1%) | 932 (30.2%) | 1,062 (38.0%) |
| Invasive Mechanical ventilation<sup>&</sup> | 2,100 (64.8%) | 491 (53.5%) | 901 (46.8%) | 346 (46.1%) | 1,473 (57.3%) | 1,491 (60.1%) |
| **1-year mortality** | **884 (24.5%)** | **341 (31.4%)** | **267 (10.1%)** | **213 (22.9%)** | **467 (15.1%)** | **657 (23.5%)** |

Data are presented as median (interquartile range), mean ± standard deviation, or number (percentage), as appropriate.
% Missing data: 43% ;& Missing data: 16%
**Abbreviations:** BMI, body mass index; CKD, chronic kidney disease; COPD, chronic obstructive pulmonary disease; ICU, intensive care unit; MCH, mean corpuscular hemoglobin; MCHC, mean
corpuscular hemoglobin concentration; MCV, mean corpuscular volume; OASIS, Oxford Acute Severity of Illness Score; PRBC, packed red blood cells; RDW, red cell distribution width; RBC, red blood
cell; SOFA, Sequential Organ Failure Assessment

### RBC Characteristics

Pre-hospital discharge RBC indices were collected a median of 9 days (IQR 5-16) after hospital admission, and 12 hours (IQR 9-30) before discharge. Median hemoglobin was 10.0 g/dL (IQR, 8.8–11.6 g/dL), and 17,247 (85.2%) patients were anemic at hospital discharge (eFigure 1). Among 8,929 females, median hemoglobin was 9.8 g/dL (IQR, 8.7–11.2 g/dL), and 7,642 females (85.6%) were anemic (eFigure 2). Among 11,304 males, median hemoglobin was 10.2 g/dL (IQR, 8.9–12.0), and 9,605 males (85.0%) were anemic (eFigure 2). Median MCV was 92.0 fL (IQR, 88.0–96.0 fL), with 17,829/20,209 (88.2%) values within the range 80.0–100.0 fL. The median MCH was 30.0 pg (IQR, 28.6–31.3 pg), with 18,863/20,208 values (93.3%) within the range 25.0–34.0 pg. Median MCHC was 32.5 g/dL (IQR, 31.5–33.5 g/dL), with 15,008/20,209 values (74.3%) within the range 31.5–35.5 g/dL. Median predischarge RBC count was 3.3 × 10¹²/L (IQR, 2.9–3.9 × 10¹²/L), with fewer than half of individuals (8,644/20,209; 42.8%) within the range 3.5–5.5 × 10¹²/L. Finally, median RDW was 14.8% (IQR, 13.6–16.5%), with 12,678/20,195 (62.8%) values within the range 11.5–15.5% (eFigure 1).

**Figure 2.**
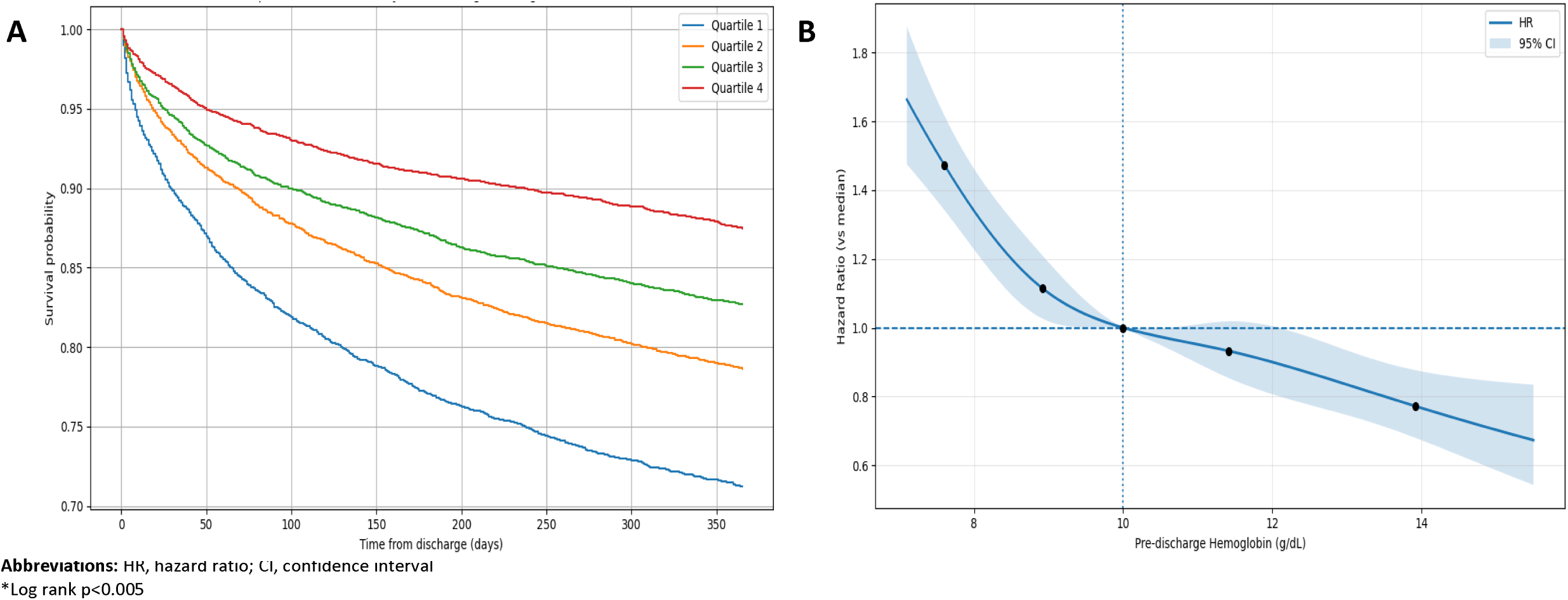
Survival and adjusted relative hazards of one-year mortality according to pre-discharge hemoglobin after critical illness. (A) Kaplan–Meier survival curves restricted cubic spline (B) Adjusted. (**A**) Kaplan–Meier survival curves showing all-cause mortality within 365 days following hospital discharge survival by pre-discharge Hemoglobin, across quartiles. Red denotes the highest quartile, followed by green and orange, then blue as the lowest quartile. Difference between groups was assessed using the log-rank test, and was statistically significant (p < 0.05). (**B**) Restricted cubic spline of adjusted hazard ratio (95% CI) for all-cause mortality across continuous pre-discharge hemoglobin, relative to the median hemoglobin of 10.0 g/dL(reference). The solid line indicates the adjusted hazard ratio and the shaded area the 95% confidence interval. The dashed horizontal line indicates unity (HR of 1.0), and the dashed vertical line indicates the median. Circles indicate model knots (5%, 27.5%, 50%, 72.5%, 95%).

### Association between Pre-Discharge Anemia and One-Year Mortality

Patients with anemia were at higher risk for one-year mortality than those without anemia (3,728 /17,247 [21.6%] versus 314/2,986 [10.5%]; adjusted HR 1.42, 95% CI 1.26–1.61; p<0.001). Anemia was associated with higher post-hospital mortality among both women (adjusted HR 1.32, 95% CI 1.12–1.56; p<0.005) and men (adjusted HR 1.56, 95% CI 1.31–1.86; p<0.001).

Overall, the frequency of post-hospital mortality demonstrated a graded relationship with pre-discharge hemoglobin (from lowest to highest hemoglobin quartile: 28.8%, 21.4%, 17.3%, and 12.5%; log-rank P<0.005; Figure 2A). Similar findings were observed in sex-stratified sensitivity analysis (eFigure 3). Adjusted hazard for post-hospital mortality was observed across the range of hemoglobin (Figure 2B).

**Figure 3.**
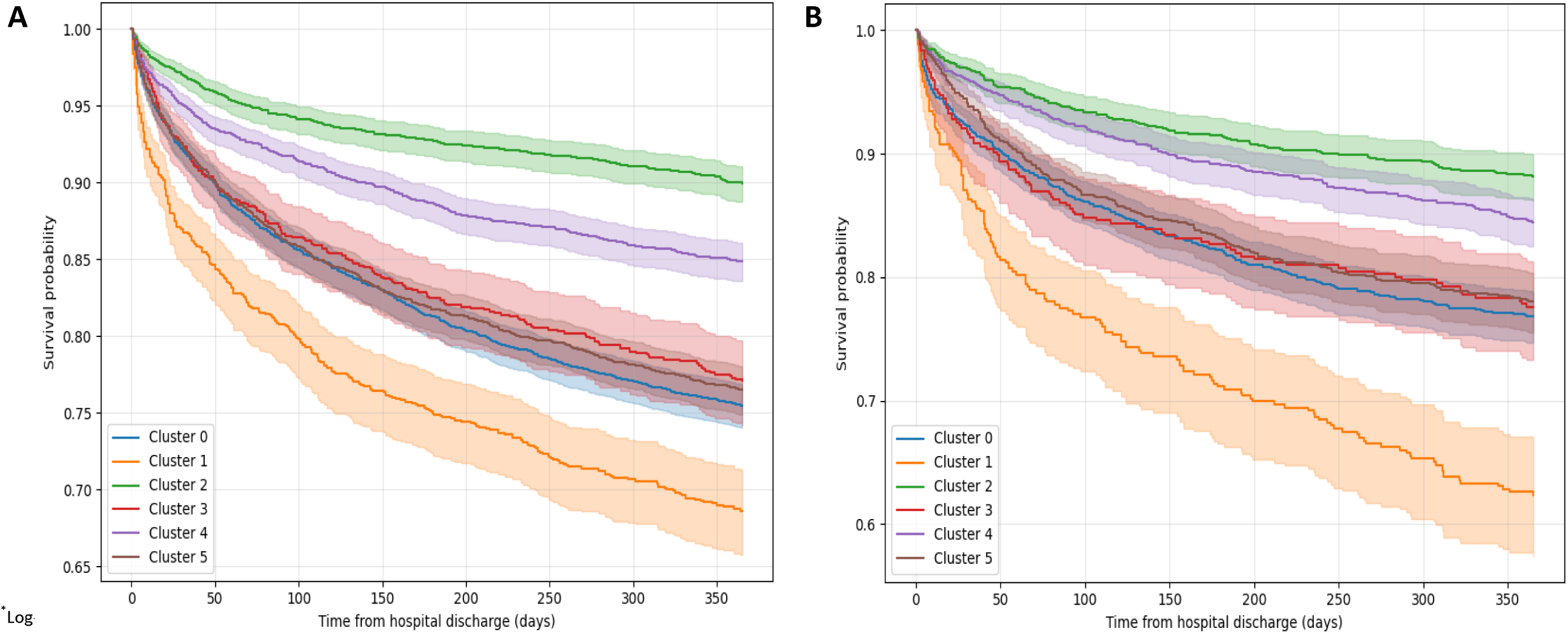
Survival after critical illness discharge according to hematologic clusters derived from RBC indices at discharge among critical illness survivors. (A) Derivation cohort. (B) Internal validation cohort. Kaplan–Meier survival curves showing one-year all-cause mortality following hospital discharge, stratified by Gaussian mixture model–derived pre-discharge RBC and erythropoiesis-related clusters. The derivation cohort is shown in the left panel (**A**), and the validation cohort is shown in the right panel (**B**). Shaded areas represent 95% confidence intervals. Time zero corresponds to hospital discharge. Differences in survival across clusters were assessed using log-rank tests and were statistically significant in both cohorts (p < 0.005)

### Clustering and Characterization of Hematologic Clusters Derived from RBC Indices

Based on the lowest BIC (eFigure 4), a six-cluster solution was selected in the derivation cohort (eFigure 5A), and this was replicated in the validation cohort (eFigure 5B). To assess the stability of the six-cluster solution, we performed bootstrap resampling, which showed consistent reproduction of the cluster structure across resampled datasets (median adjusted Rand index=0.98). Although the Silhouette coefficient was 0.078 and the Davies–Bouldin index was 2.74, indicating limited separation and overlap among the identified clusters, we retained the six-cluster solution because this exploratory cluster analysis aimed to identify more granular and potentially clinically meaningful patterns across multiple RBC indices rather than define mutually exclusive disease categories. Accordingly, the clusters should be interpreted as hypothesis-generating and require external validation.

Baseline demographic and clinical characteristics differed across the six clusters in the derivation cohort (14,163) (Table 2). Cluster distribution was: 0 (25.5%), 1 (7.7%), 2 (18.8%), 3 (6.6%), 4 (21.8%), and 5 (19.7%). Clusters 0 and 3 were comprised of patients with a higher burden of chronic comorbidities, including cardiovascular, cerebrovascular, and renal disease. Cluster 1 included comparatively younger patients with a high prevalence of CKD (21.4%), severe liver disease, and cancer. Acute illness severity varied across clusters: clusters 0 and 1 had higher SOFA scores, while clusters 0 and 5 more frequently required organ support, including mechanical ventilation and vasopressors. In contrast, cluster 2 had the lowest illness severity, shorter ICU and hospital stays, and minimal organ support. Cluster 4 was characterized by older age and a higher proportion of surgical procedures. Similar patterns were observed in the validation cohort (eTable 2).

Association between Hematologic Clusters Derived from RBC Indices and One-Year Mortality In the derivation cohort, 20.0% of patients died within 365 days of discharge, with a median time from hospital discharge to death of 69 (IQR 19–174) days. Mortality across clusters ranged from 10.1% in cluster 2 to 31.4% in cluster 1, with intermediate-lower mortality in cluster 4 (15.1%) and intermediate-higher mortality in clusters 3 (22.9%), 5 (23.5%), and 0 (24.5%). Kaplan–Meier analyses demonstrated clear separation in post-discharge mortality across clusters (log-rank p<0.005, Figure 3A). Results were similar in the validation cohort (log-rank p<0.005, Figure 3B).

The proportional hazards assumption for the cluster terms was satisfied in the multivariable-adjusted models in both the derivation and validation cohorts. A graded increase in long-term mortality risk across hematologic clusters derived from RBC indices persisted after multivariable adjustment (eTable 3). Using cluster 2 as the reference (lowest risk cluster), the highest mortality risk was observed in the cluster characterized by elevated RDW and MCV (cluster 1; derivation cohort adjusted HR 2.14, 95% CI 1.81-2.53). Adjusted mortality associations for the remaining clusters were intermediate to these groups, and findings were consistent in the validation population (eTable 3).

## Discussion

In this retrospective cohort of more than 20,000 adult ICU survivors, lower pre-discharge hemoglobin was associated with one-year post-discharge mortality. Data-driven clustering of pre-discharge RBC indices identified six hematologic clusters with differing patterns of anemia of putatively differing etiologies. Those clusters characterized by persistent anemia and elevated RDW and MCV had the highest one-year mortality, whereas patients with normal RDW and otherwise preserved RBC indices had lower risk. Collectively, these findings suggest that anemia and hematologic clusters derived from routinely available pre-discharge RBC indices may help refine post-discharge one-year mortality risk assessment and provide hypothesis-generating insights into causes of anemia and RBC dysfunction after critical illness.

There is an unmet clinical need to improve long-term survival and to reduce disability following critical illness. Studies evaluating prognostic biomarkers and potential mediators of mortality risk are needed. Anemia and distinct RBC-index derangements were associated with mortality and may represent prognostic biomarkers. Few studies to date have evaluated pre-hospital discharge prognostic biomarkers, including RBC indices and long-term post-hospital outcomes.[17, 18] Pre-discharge RBC indices may reflect chronic health state and underlying comorbidities or putatively capture persistent erythropoiesis disturbances following critical illness, driven by sustained inflammation,[6] impaired iron metabolism,[19] oxidative stress,[20] nutritional deficiencies,[21] or clonal hematopoiesis associated with somatic mutations.[22]

Anemia at discharge has been associated with 90-day rehospitalization and mortality in a prior cohort of predominantly male sepsis survivors.[17] Our study supports pre-discharge hemoglobin as a potential prognostic biomarker after critical illness and extends prior work by integrating multiple RBC indices into an analytic framework. Furthermore, ongoing anemia may putatively contribute to persistent functional limitation post-hospital. In patients with chronic heart failure and chronic renal disease, correcting anemia improves outcomes. The present results are hypothesis-generating and cannot guide treatment decisions, but do encourage future study.

Because RBC indices reflect overlapping but non-redundant aspects of nutritional status, chronic disease, inflammation, and marrow function, joint profiling may better capture the underlying biological states than individual parameters. These biological processes likely span critical illness syndromes, and clusters may provide insights to support evidence-based precision medicine.[10] Previous studies have shown that unsupervised machine learning applied to admission data can identify distinct clusters with differential risk for in-hospital mortality among critically ill patients with heart failure,[23] acute respiratory distress syndrome,[24] and sepsis.[25] In our analysis, hematologic clusters derived from RBC indices capture erythroid heterogeneity beyond isolated parameters.

Routine RBC indices may provide an inexpensive and widely available data that identifies critical illness survivors at higher post-discharge mortality risk. Low MCV patterns with elevated RDW— observed in cluster 3—have been linked to iron-restricted erythropoiesis and adverse outcomes among cardiac ICU patients.[26] Elevated MCV and RDW, characteristic of the cluster with highest one-year mortality, have been associated with increased risk, even in non–critically ill populations.[27] This cluster may reflect acute inflammatory suppression of erythroid maturation or stress erythropoiesis during severe systemic illness. In contrast, clusters with preserved hemoglobin, normal RDW, and stable RBC indices —characteristic of the clusters with the lowest one-year mortality —likely reflect recovery of hematopoietic homeostasis.

Several limitations merit consideration. First, pre-discharge RBC indices may be influenced by inpatient therapies, physician practice, and timing of assessment, which our study cannot completely capture. However, RBC indices were predominantly measured within 24 hours of discharge and were available for more than 90% of the cohort, mitigating the risk of residual confounding. Second, the observational design precluded access to other key related biomarkers, including hematinic factor profiles (e.g. iron, B12) and reticulocyte indices, limiting diagnosis of causes of anemia. Nevertheless, given the complexity of erythropoietic regulation and the limited routine availability of these biomarkers, reliance on RBC indices enhances clinical applicability. Third, the single-center design may limit generalizability. Although findings were consistent in internal validation, external validation in independent cohorts is needed.

Fourth, reliance on state rather than national mortality data may have led to nonrandom underascertainment of out-of-hospital deaths; however, the observed one-year mortality rate was within the expected range for a general critically ill population. Finally, our study is exploratory; future studies should incorporate additional clinical and biological variables and validate these findings in multicenter cohorts.

## Conclusion

In this large cohort of ICU survivors, both pre-discharge anemia and hematologic clusters derived from RBC indices were associated with one-year post-discharge mortality. These hypothesis-generating findings support further evaluation of pre-discharge RBC indices as an inexpensive, readily available, and pragmatic framework for post–critical illness risk stratification and future interventional studies. External validation is required to confirm these results.

## List of abbreviations

CBC: Complete blood count
CI: Confidence interval
GMM: Gaussian mixture model
HR: Hazard ratio
ICD: International Classification of Diseases
ICU: Intensive care unit
IQR: Interquartile range
MCV: Mean corpuscular volume
MCH: Mean corpuscular hemoglobin
MCHC: Mean corpuscular hemoglobin concentration
MICE: Multiple imputation by chained equations
MIMIC-IV: Medical Information Mart for Intensive Care
IV OASIS: Oxford Acute Severity of Illness Score
RBC: Red blood cell
RDW: Red cell distribution width
SOFA: Sequential Organ Failure Assessment
WHO: World Health Organization

## Declarations

### Ethics approval and consent to participate

MIMIC-IV was developed with approval from the institutional review boards of the Massachusetts Institute of Technology and BIDMC (IRB #2001P001699), with a waiver of informed consent due to data de-identification. The study author (L.M.) completed the required training and was authorized to access the database. This study has also obtained local ethics approval from the McGill University Health Centre Research Ethics Board (2026-12503).

### Consent for publication

Not applicable

### Availability of data and materials

The datasets used and/or analyzed during the current study are available from the corresponding author on reasonable request.

### Authors’ contributions

LM performed the statistical analysis. LM, DJ, and PRL were the major contributors to writing the manuscript. The study was designed by LM, PRL, RZ, JM, and DJ. All authors critically appraised the data. All authors read and approved the final manuscript.

## Data Availability

All data produced in the present study are available upon reasonable request to the authors

https://physionet.org/content/mimiciv/3.1/

## Acknowledgements

PRL is supported by a career award from the Fonds de recherche du Quebec.

## References

1. Lone NI, Gillies MA, Haddow C, Dobbie R, Rowan KM, Wild SH, et al. Five-Year Mortality and Hospital Costs Associated with Surviving Intensive Care. Am J Respir Crit Care Med. 2016;194(2):198–208. doi: 10.1164/rccm.201511-2234OC.

2. Doherty Z, Kippen R, Bevan D, Duke G, Williams S, Wilson A, et al. Long-term outcomes of hospital survivors following an ICU stay: A multi-centre retrospective cohort study. PLoS One. 2022;17(3):e0266038. doi: 10.1371/journal.pone.0266038.

3. Jentzer JC, Lawler PR, Van Houten HK, Yao X, Kashani KB, Dunlay SM. Cardiovascular Events Among Survivors of Sepsis Hospitalization: A Retrospective Cohort Analysis. J Am Heart Assoc. 2023;12(3):e027813. doi: 10.1161/JAHA.122.027813.

4. Kosyakovsky LB, Angriman F, Katz E, Adhikari NK, Godoy LC, Marshall JC, et al. Association between sepsis survivorship and long-term cardiovascular outcomes in adults: a systematic review and meta-analysis. Intensive Care Med. 2021;47(9):931–42. doi: 10.1007/s00134-021-06479-y.

5. Salvagno GL, Sanchis-Gomar F, Picanza A, Lippi G. Red blood cell distribution width: A simple parameter with multiple clinical applications. Crit Rev Clin Lab Sci. 2015;52(2):86–105. doi: 10.3109/10408363.2014.992064.

6. Lippi G, Targher G, Montagnana M, Salvagno GL, Zoppini G, Guidi GC. Relation between red blood cell distribution width and inflammatory biomarkers in a large cohort of unselected outpatients. Arch Pathol Lab Med. 2009;133(4):628–32. doi: 10.5858/133.4.628.

7. Mitrache C, Passweg JR, Libura J, Petrikkos L, Seiler WO, Gratwohl A, et al. Anemia: an indicator for malnutrition in the elderly. Ann Hematol. 2001;80(5):295–8. doi: 10.1007/s002770100287.

8. Maddox TM, Januzzi JL, Jr., Allen LA, Breathett K, Brouse S, Butler J, et al. 2024 ACC Expert Consensus Decision Pathway for Treatment of Heart Failure With Reduced Ejection Fraction: A Report of the American College of Cardiology Solution Set Oversight Committee. J Am Coll Cardiol. 2024;83(15):1444-88. doi: 10.1016/j.jacc.2023.12.024.

9. Kidney Disease: Improving Global Outcomes Anemia Work G. KDIGO 2026 Clinical Practice Guideline for the Management of Anemia in Chronic Kidney Disease (CKD). Kidney Int. 2026;109(1S):S1–S99. doi: 10.1016/j.kint.2025.06.006.

10. Gordon AC, Alipanah-Lechner N, Bos LD, Dianti J, Diaz JV, Finfer S, et al. From ICU Syndromes to ICU Subphenotypes: Consensus Report and Recommendations for Developing Precision Medicine in the ICU. Am J Respir Crit Care Med. 2024;210(2):155–66. doi: 10.1164/rccm.202311-2086SO.

11. Johnson AEW, Bulgarelli L, Shen L, Gayles A, Shammout A, Horng S, et al. MIMIC-IV, a freely accessible electronic health record dataset. Sci Data. 2023;10(1):1. doi: 10.1038/s41597-022-01899-x.

12. Thygesen SK, Christiansen CF, Christensen S, Lash TL, Sorensen HT. The predictive value of ICD-10 diagnostic coding used to assess Charlson comorbidity index conditions in the population-based Danish National Registry of Patients. BMC Med Res Methodol. 2011;11:83. doi: 10.1186/1471-2288-11-83.

13. Huang Y, Yang Z, Rahmani A. MIMIC-Sepsis: A Curated Benchmark for Modeling and Learning from Sepsis Trajectories in the ICU. 2025 doi: 10.48550/arXiv.2510.24500.

14. Guideline on haemoglobin cutoffs to define anaemia in individuals and populations. WHO Guidelines Approved by the Guidelines Review Committee. Geneva2024.

15. Austin PC, Fang J, Lee DS. Using fractional polynomials and restricted cubic splines to model non-proportional hazards or time-varying covariate effects in the Cox regression model. Stat Med. 2022;41(3):612–24. doi: 10.1002/sim.9259.

16. Harrell FE. Regression Modeling Strategies. 2nd ed. Springer New York, NY; 2010.

17. Denstaedt SJ, Cano J, Wang XQ, Donnelly JP, Seelye S, Prescott HC. Blood count derangements after sepsis and association with post-hospital outcomes. Front Immunol. 2023;14:1133351. doi: 10.3389/fimmu.2023.1133351.

18. Soussi S, Sharma D, Juni P, Lebovic G, Brochard L, Marshall JC, et al. Identifying clinical subtypes in sepsis-survivors with different one-year outcomes: a secondary latent class analysis of the FROG-ICU cohort. Crit Care. 2022;26(1):114. doi: 10.1186/s13054-022-03972-8.

19. Allen LA, Felker GM, Mehra MR, Chiong JR, Dunlap SH, Ghali JK, et al. Validation and potential mechanisms of red cell distribution width as a prognostic marker in heart failure. J Card Fail. 2010;16(3):230–8. doi: 10.1016/j.cardfail.2009.11.003.

20. Joosse HJ, van Oirschot BA, Kooijmans SAA, Hoefer IE, van Wijk RAH, Huisman A, et al. In-vitro and in-silico evidence for oxidative stress as drivers for RDW. Sci Rep. 2023;13(1):9223. doi: 10.1038/s41598-023-36514-5.

21. Haenggi E, Kaegi-Braun N, Wunderle C, Tribolet P, Mueller B, Stanga Z, et al. Red blood cell distribution width (RDW) - A new nutritional biomarker to assess nutritional risk and response to nutritional therapy? Clin Nutr. 2024;43(2):575–85. doi: 10.1016/j.clnu.2024.01.001.

22. Kar SP, Quiros PM, Gu M, Jiang T, Mitchell J, Langdon R, et al. Genome-wide analyses of 200,453 individuals yield new insights into the causes and consequences of clonal hematopoiesis. Nat Genet. 2022;54(8):1155–66. doi: 10.1038/s41588-022-01121-z.

23. Jentzer JC, Reddy YNV, Soussi S, Crespo-Diaz R, Patel PC, Lawler PR, et al. Unsupervised machine learning to identify subphenotypes among cardiac intensive care unit patients with heart failure. ESC Heart Fail. 2024;11(6):4242–56. doi: 10.1002/ehf2.15027.

24. Sinha P, Delucchi KL, McAuley DF, O’Kane CM, Matthay MA, Calfee CS. Development and validation of parsimonious algorithms to classify acute respiratory distress syndrome phenotypes: a secondary analysis of randomised controlled trials. Lancet Respir Med. 2020;8(3):247–57. doi: 10.1016/S2213-2600(19)30369-8.

25. Kudo D, Goto T, Uchimido R, Hayakawa M, Yamakawa K, Abe T, et al. Coagulation phenotypes in sepsis and effects of recombinant human thrombomodulin: an analysis of three multicentre observational studies. Crit Care. 2021;25(1):114. doi: 10.1186/s13054-021-03541-5.

26. Luo F, Wang Z, Gao T, Wang B, Gao Y, Liu M, et al. Impact of Hemoglobin and Iron Deficiency on Mortality in Patients with Acute Myocardial Infarction in Intensive Care Units: A Retrospective Study from MIMIC-IV. Rev Cardiovasc Med. 2025;26(5):28261. doi: 10.31083/RCM28261.

27. Lam AP, Gundabolu K, Sridharan A, Jain R, Msaouel P, Chrysofakis G, et al. Multiplicative interaction between mean corpuscular volume and red cell distribution width in predicting mortality of elderly patients with and without anemia. Am J Hematol. 2013;88(11):E245–9. doi: 10.1002/ajh.23529.

