## Supplemental appendix for "Association between Anemia and Hematologic Traits at Hospital Discharge with One-Year Mortality Among Survivors of Critical Illness"

**Table of Contents**

**Supplemental Figures and Tables**

eFigure 1. Distribution of pre-discharge RBC indices………....…………….…………………………..…….....…3

eFigure 2. Distribution of pre-discharge Hemoglobin by sex………....…………….…………….......….....…4

eFigure 3. Kaplan–Meier survival curves according to pre-discharge hemoglobin quartiles after critical illness. (A) Female cohort (B) Male cohort………....…..............……………….................…………5

eFigure 4. Bayesian information criterion across gaussian mixture model components in the training cohort ………....….............……........................................…….………………….................…………6

eFigure 5. Visualization of hematologic clusters derived from RBC indices at discharge among critical illness survivors. (A) Derivation cohort. (B) Internal validation cohort ………...............................................................................................................................……7

eTable 1. Baseline characteristics of critical illness survivors in derivation and validation cohorts

................…................................................................................................................................…...8

eTable 2. Baseline characteristics by hematologic clusters derived from RBC indices in the validation cohort........................................................................................................….....…....9

eTable 3. Hematologic clusters derived from RBC indices at hospital discharge and association with one-year mortality...............................................................................….....…....10

**Supplemental figures legends**

**eFigure 1. Distribution of pre-discharge RBC indices**
**(A)** Histograms and density plots showing distributions of hemoglobin, RDW, MCV, MCH, MCHC, and RBC count measured within 72 hours prior to hospital discharge in the total cohort.

**(B)** Histograms and density plots showing distributions of hemoglobin, RDW, MCV, MCH, MCHC, and RBC count measured within 72 hours prior to hospital discharge in the derivation (TRAIN) and validation (TEST) cohort.

**eFigure 2. Distribution of pre-discharge Hemoglobin by sex**

Histograms and density plots show the distribution of hemoglobin by sex, measured within 72 hours before hospital discharge. The left panel shows the female cohort (n=8,929), with the dashed blue line indicating the World Health Organization anemia cut-off for nonpregnant women (12 g/dL). The right panel shows the male cohort (n=11,304), with the dashed blue line indicating the World Health Organization anemia cut-off for men (13 g/dL).

**eFigure 3. Kaplan–Meier survival curves according to pre-discharge hemoglobin quartiles after critical illness. (A) Female cohort (B) Male cohort**
Kaplan–Meier survival curves showing all-cause mortality within 365 days following hospital discharge survival by pre-discharge Hemoglobin, across quartiles. Red denotes the highest quartile, followed by green and orange, then blue as the lowest quartile. Differences between groups were assessed using the log-rank test, and were statistically significant in both female and male cohorts (p < 0.05).

**eFigure 4. Bayesian information criterion across gaussian mixture model components in the training cohort**

Bayesian information criterion (BIC) values for Gaussian mixture models with one to six components. Lower BIC values indicate better model fit while accounting for model complexity; the six-component solution had the lowest BIC and was selected for subsequent analyses.

**eFigure 5. Visualization of clusters based on hematologic clusters derived from RBC indices at discharge among critical illness survivors. (A) Derivation cohort. (B) Internal validation cohort**

Gaussian mixture model was used to generate clusters based on predischarge RBC indices (hemoglobin, MCV, MCH, MCHC, RDW, and RBC count), with two-dimensional t-distributed stochastic neighbor embedding (t-SNE) projections showing clusters by RBC and erythropoiesis-related clusters.

**(A)** t-SNE visualization of the derivation cohort, with six distinct clusters.

**(B)** t-SNE visualization of the validation cohort, reproducing cluster assignments.

Each point represents a single patient, and the colors indicate assigned cluster. Axes represent t-SNE dimensions and have no direct clinical interpretation. Similar spatial separation and cluster structure across cohorts demonstrate preservation of erythropoiesis clusters in the validation cohort.

**eFigure 1. Distribution of pre-discharge RBC indices**
**A**

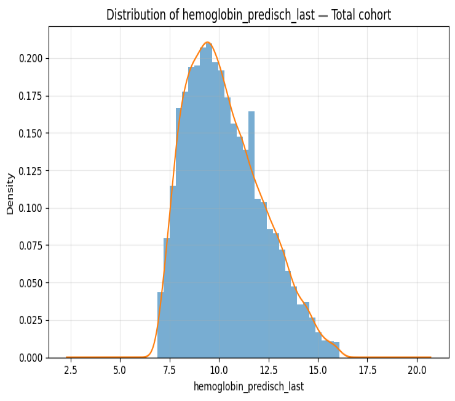

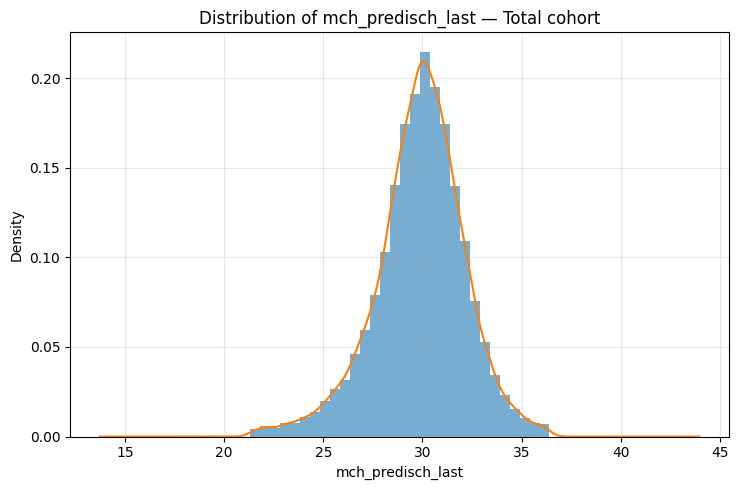

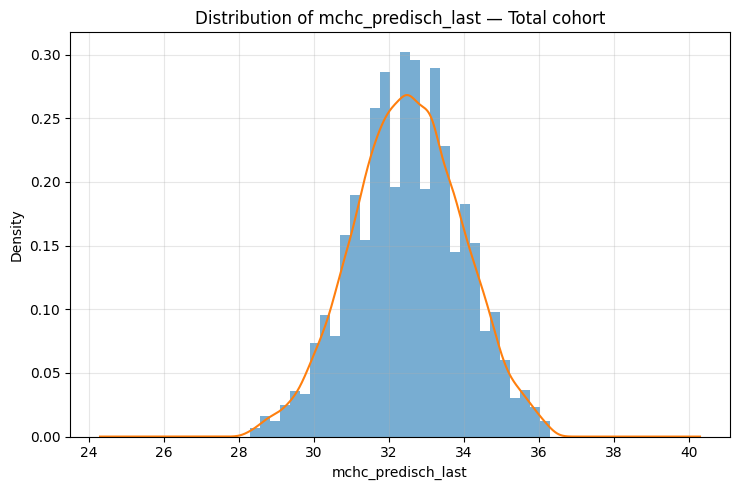

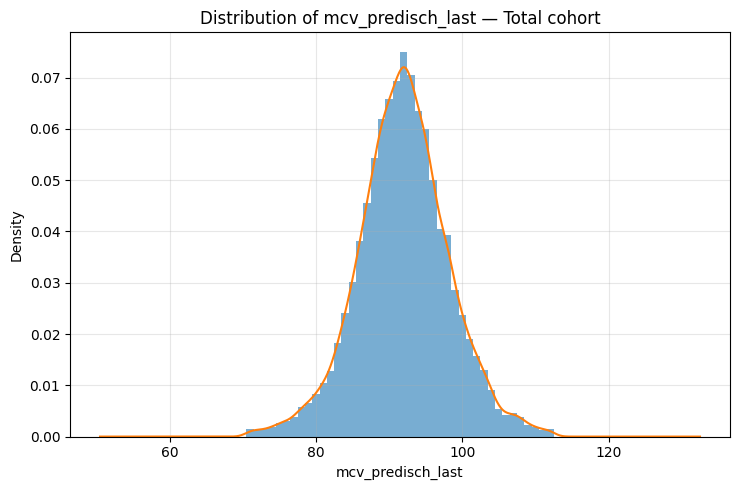

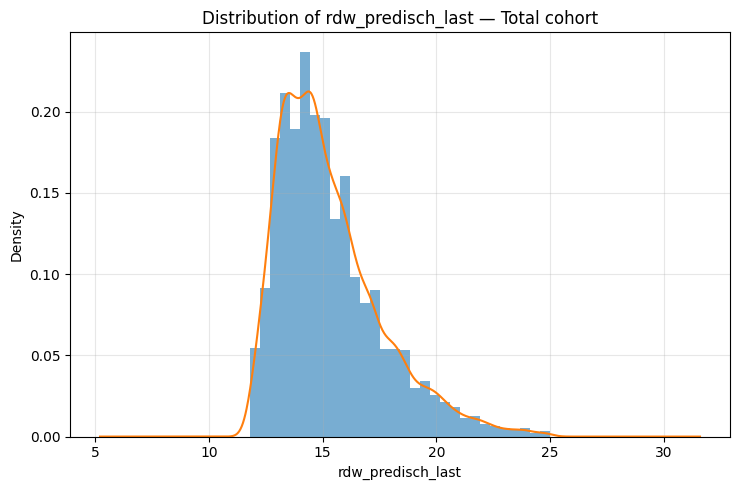

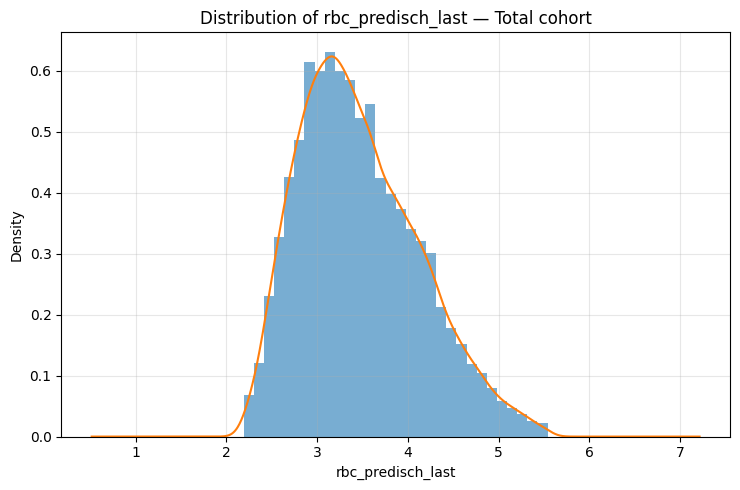

**B**

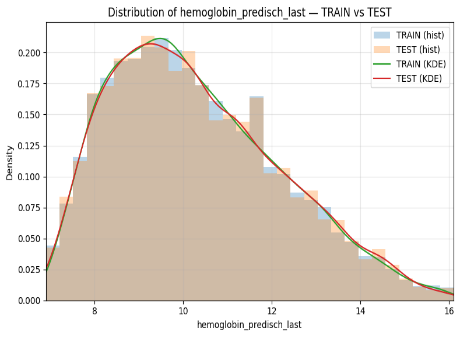

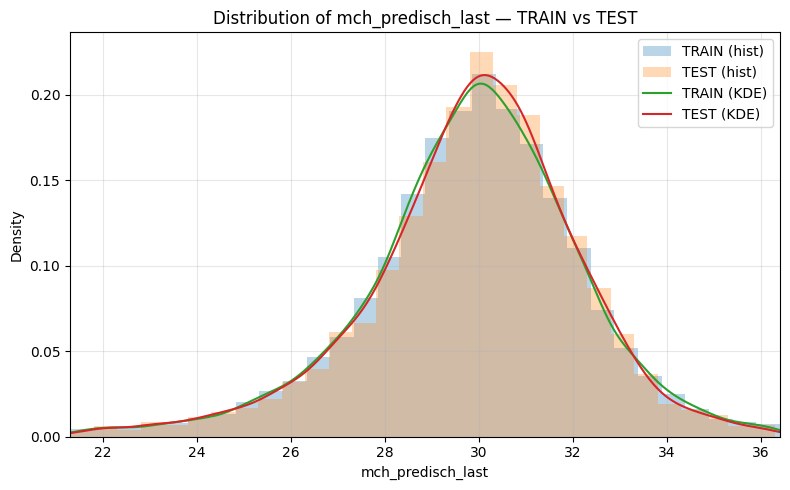

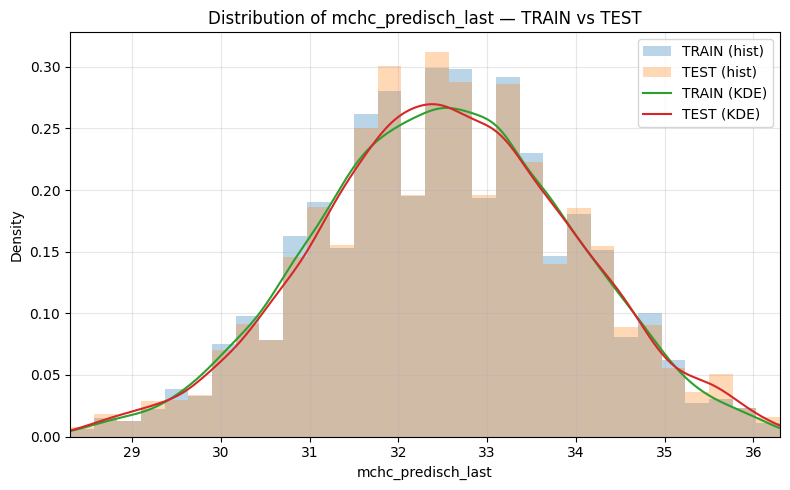

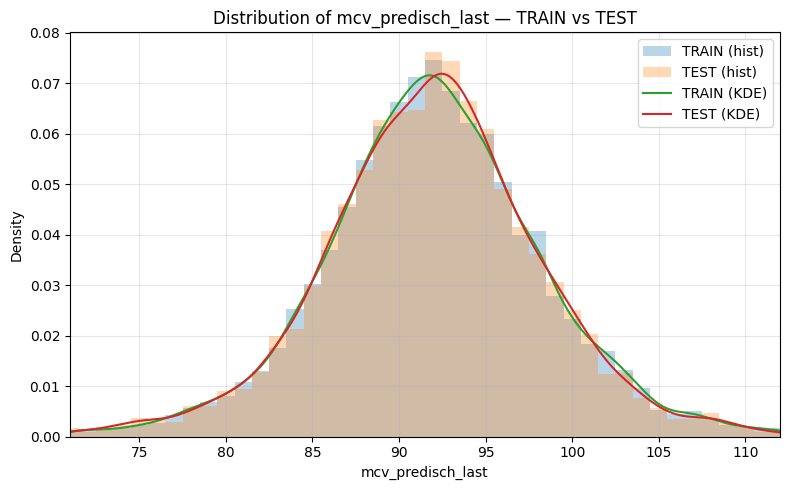

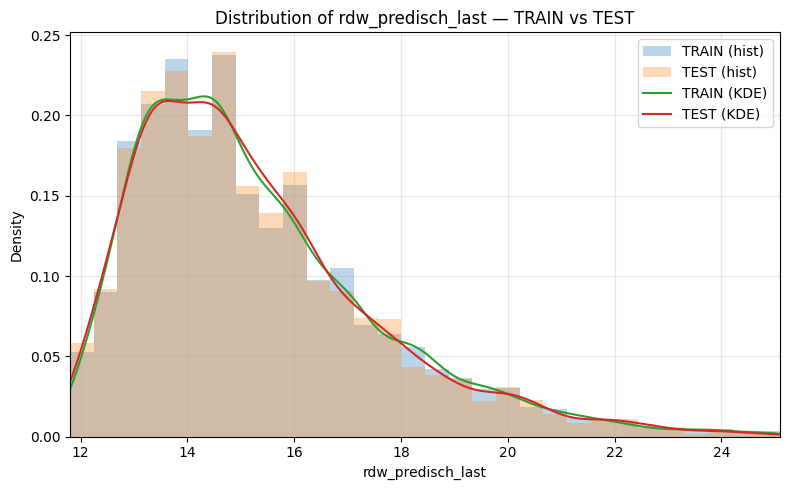

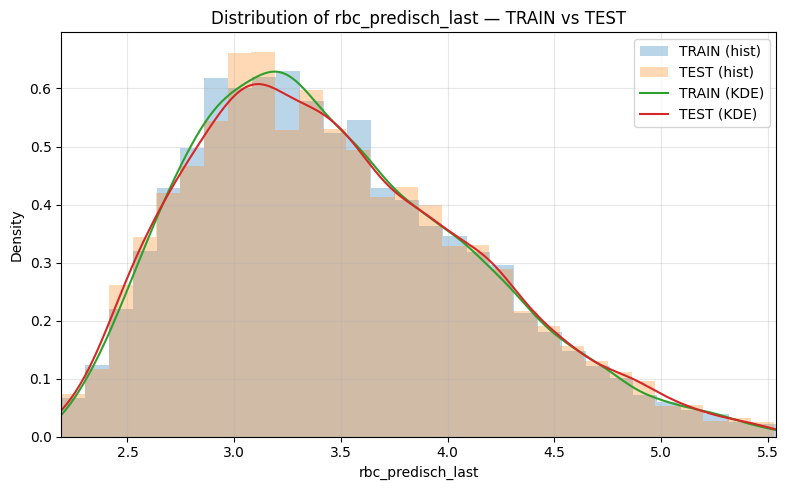

**Abbreviations:** Hb, hemoglobin; MCH, mean corpuscular hemoglobin; MCHC, mean corpuscular hemoglobin concentration; MCV, mean corpuscular volume; RDW, red cell distribution width; RBC, red blood cell.

**eFigure 2. Distribution of pre-discharge Hemoglobin by sex**

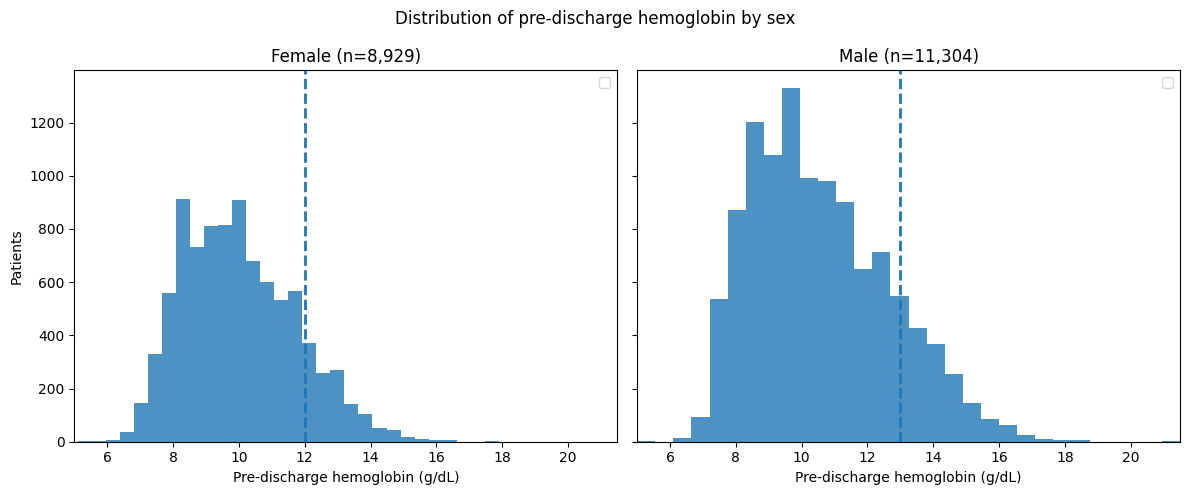

**A**

**B**

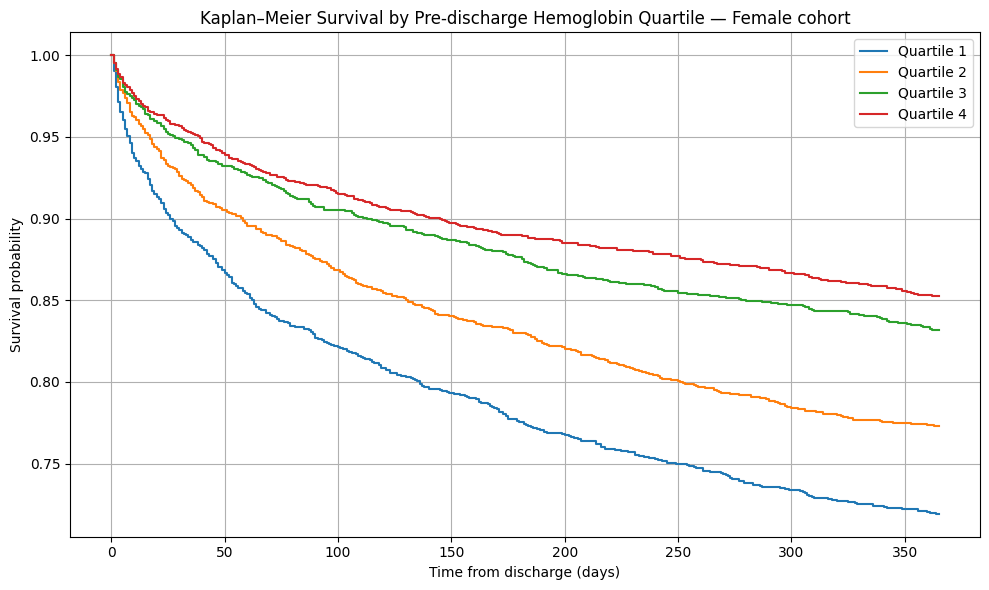

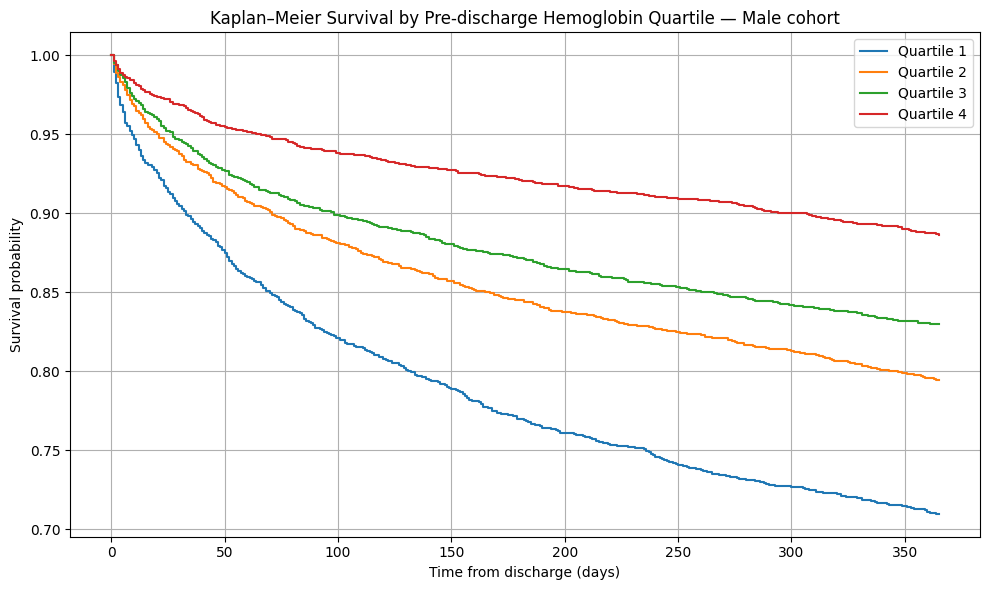

**eFigure 3. Kaplan–Meier survival curves according to pre-discharge hemoglobin quartiles after critical illness. (A) Female cohort (B) Male cohort**

**eFigure 4. Bayesian information criterion across gaussian mixture model components in the training cohort**

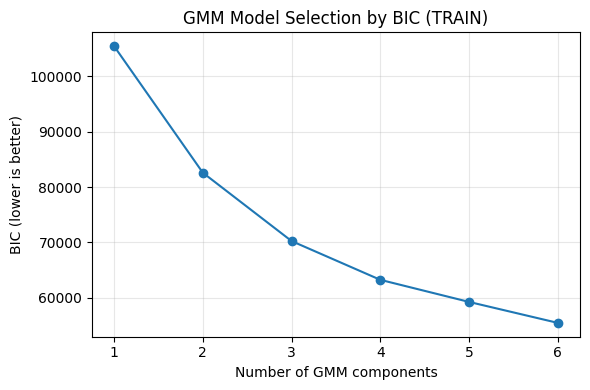

**eFigure 5. Visualization of hematologic clusters derived from RBC indices at discharge among critical illness survivors. (A) Derivation cohort. (B) Internal validation cohort**

**B**

**A**

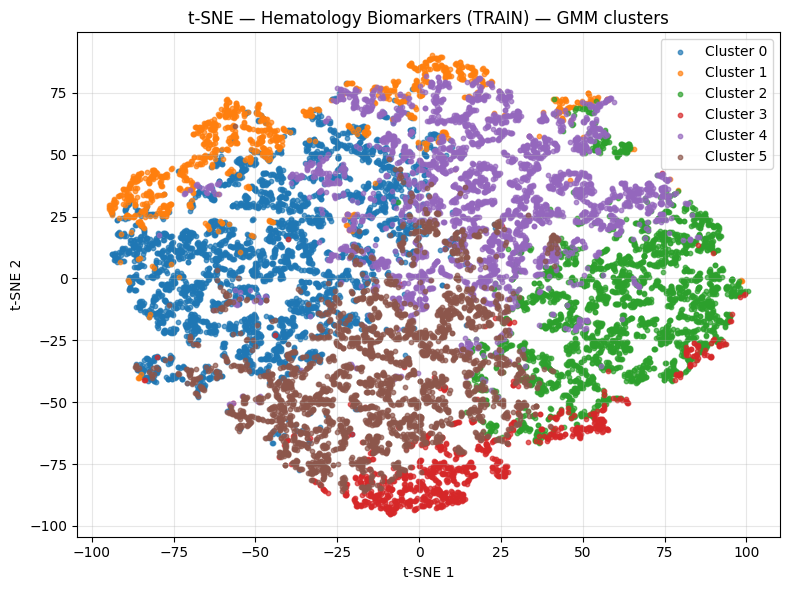

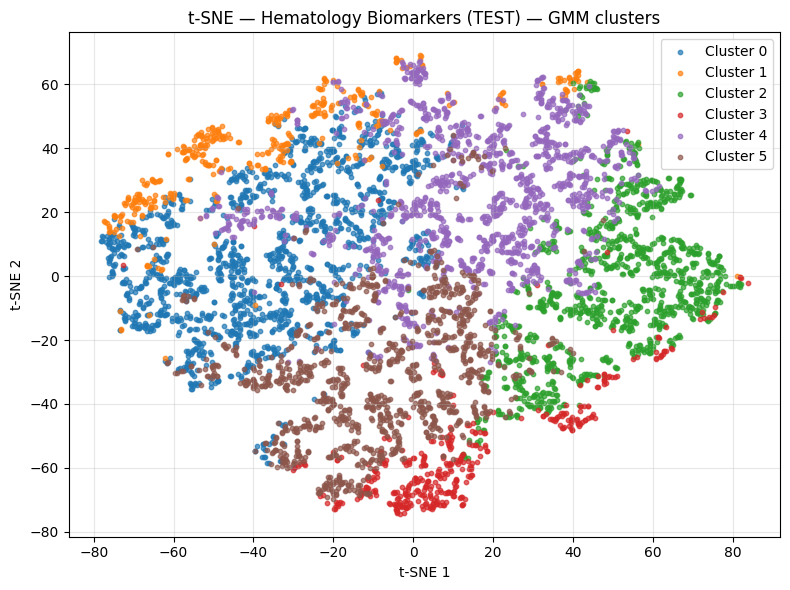

**eTable 1. Baseline characteristics of critical illness survivors in derivation and validation cohorts**

| Characteristic | Derivation Cohort  (n = 14,163) | Validation Cohort  (n = 6,071) |
| --- | --- | --- |
| ****Pre-discharge indices**** |  |  |
| Hemoglobin, g/dL | 10.00 (8.80–11.60) | 10.00 (8.80–11.60) |
| RDW, % | 14.80 (13.60–16.50) | 14.80 (13.60–16.40) |
| MCV, fL | 92.00 (88.00–96.00) | 92.00 (88.00–96.00) |
| MCH, pg | 30.00 (28.60–31.30) | 30.00 (28.70–31.30) |
| MCHC, g/dL | 32.50 (31.50–33.50) | 32.50 (31.50–33.50) |
| RBC, ×10¹²/L | 3.37 (2.97–3.90) | 3.38 (2.96–3.92) |
| ****Demographics**** |  |  |
| Age, years | 66.00 (54.00–78.00) | 67.00 (55.00–78.00) |
| Male sex | 7,897 (55.8%) | 3,408 (56.1%) |
| BMI, kg/m²^%^ | 28.07 (24.30–33.06) | 28.05 (24.34–32.63) |
| ****Comorbidities**** |  |  |
| Charlson index | 5.00 (3.00–7.00) | 5.00 (3.00–7.00) |
| Myocardial infarction | 2,605 (18.4%) | 1,195 (19.7%) |
| Heart failure | 4,164 (29.4%) | 1,794 (29.6%) |
| Peripheral vascular disease | 1,550 (10.9%) | 746 (12.3%) |
| Cerebrovascular disease | 2,969 (21.0%) | 1,287 (21.2%) |
| COPD | 3,406 (24.0%) | 1,475 (24.3%) |
| Diabetes mellitus | 3,378 (23.9%) | 1,428 (23.5%) |
| Chronic kidney disease | 2,809 (19.8%) | 1,235 (20.3%) |
| Malignancy | 1,647 (11.6%) | 651 (10.7%) |
| ****Critical illness and severity**** |  |  |
| SOFA score (24 h) | 4.00 (2.00–7.00) | 4.00 (2.00–7.00) |
| OASIS score | 32.00 (26.00–37.00) | 32.00 (26.00–37.00) |
| ICU length of stay, days | 3.75 (2.67–6.33) | 3.79 (2.67–6.33) |
| Hospital length of stay, days | 10.00 (6.00–16.00) | 10.00 (6.00–16.00) |
| Sepsis (Sepsis-3) | 6,233 (44.0%) | 2,680 (44.1%) |
| ****In-hospital therapies**** |  |  |
| Surgery | 7,424 (52.4%) | 3,172 (52.2%) |
| PRBC transfusion | 3,369 (23.8%) | 1,467 (24.2%) |
| PRBC transfusion, units | 0.90 ± 2.67 | 0.89 ± 2.79 |
| Vasopressors use (first 48 h) | 4,535 (32.0%) | 1,956 (32.2%) |
| Invasive mechanical ventilation^&^ | 6,802 (57.2%) | 2,912 (57.1%) |

Data are presented as median (interquartile range), mean ± standard deviation, or number (percentage), as appropriate. Pre-discharge laboratory values were obtained within 72 hours prior to hospital discharge.

^%^ Missing data: 43% in both derivation and validation cohorts; ^&^ Missing data: 16% in both derivation and validation cohorts

**Abbreviations:** BMI, body mass index; COPD, chronic obstructive pulmonary disease; ICU, intensive care unit; MCH, mean corpuscular hemoglobin; MCHC, mean corpuscular hemoglobin concentration; MCV, mean corpuscular volume; OASIS, Oxford Acute Severity of Illness Score; PRBC, packed red blood cells; RDW, red cell distribution width; RBC, red blood cell; SOFA, Sequential Organ Failure Assessment.

**eTable 2. Baseline characteristics by hematologic clusters derived from RBC indices in the validation cohort**

| **Characteristic** | **Cluster 0 (n=1,587)** | **Cluster 1 (n=409)** | **Cluster 2 (n=1,158)** | **Cluster 3 (n=415)** | **Cluster 4 (n=1,366)** | **Cluster 5 (n=1,136)** |
| --- | --- | --- | --- | --- | --- | --- |
| **Pre-discharge indices** |  |  |  |  |  |  |
| Hemoglobin, g/dL | 8.70 (8.00–9.20) | 8.90 (8.00–10.20) | 12.80 (11.90–13.70) | 10.40 (9.10–12.40) | 10.90 (10.30–11.60) | 9.40 (8.60–10.10) |
| RDW, % | 15.60 (14.40–17.20) | 16.30 (14.30–19.20) | 13.60 (13.00–14.40) | 17.30 (15.20–19.60) | 14.00 (13.20–15.10) | 15.50 (14.50–17.10) |
| MCV, fL | 95.00 (92.00–97.00) | 103.00 (100.0–107.0) | 90.00 (87.00–92.00) | 80.00 (76.00–84.00) | 94.00 (91.00–96.00) | 88.00 (85.00–90.00) |
| MCH, pg | 30.40 (29.70–31.20) | 33.50 (32.70–34.70) | 29.70 (28.80–30.60) | 25.10 (23.40–26.40) | 31.10 (30.40–31.90) | 28.10 (27.20–28.80) |
| MCHC, g/dL | 32.10 (31.30–33.00) | 33.00 (31.90–33.90) | 33.10 (32.30–34.00) | 31.10 (30.10–32.00) | 33.20 (32.40–34.00) | 31.80 (31.00–32.70) |
| RBC, ×10¹²/L | 2.85 (2.67–3.00) | 2.64 (2.42–3.06) | 4.29 (4.10–4.56) | 4.22 (3.81–4.88) | 3.53 (3.34–3.73) | 3.33 (3.13–3.55) |
| **Demographics** |  |  |  |  |  |  |
| Age, years | 70.00 (58.00–79.00) | 66.00 (56.00–79.00) | 62.00 (49.00–74.00) | 64.00 (50.00–76.00) | 68.00 (57.00–79.00) | 68.00 (56.00–79.00) |
| Male sex | 877 (55.3%) | 229 (56.0%) | 768 (66.3%) | 220 (53.0%) | 743 (54.4%) | 571 (50.3%) |
| BMI, kg/m²^%^ | 27.80 (24.48–32.30) | 27.59 (23.14–32.41) | 28.39 (24.59–32.63) | 28.88 (25.07–34.70) | 27.55 (23.93–31.50) | 28.70 (24.69–34.03) |
| **Comorbidities** |  |  |  |  |  |  |
| Charlson index | 5.00 (3.00–7.00) | 5.00 (4.00–7.00) | 4.00 (2.00–6.00) | 5.00 (2.00–7.00) | 4.00 (3.00–6.00) | 5.00 (3.00–7.00) |
| Myocardial infarction | 386 (24.3%) | 57 (13.9%) | 184 (15.9%) | 72 (17.3%) | 258 (18.9%) | 238 (21.0%) |
| Heart failure | 509 (32.1%) | 125 (30.6%) | 266 (23.0%) | 148 (35.7%) | 355 (26.0%) | 391 (34.4%) |
| Peripheral vascular disease | 232 (14.6%) | 63 (15.4%) | 100 (8.6%) | 51 (12.3%) | 166 (12.2%) | 134 (11.8%) |
| Cerebrovascular disease | 257 (16.2%) | 62 (15.2%) | 353 (30.5%) | 83 (20.0%) | 336 (24.6%) | 196 (17.3%) |
| COPD | 381 (24.0%) | 105 (25.7%) | 242 (20.9%) | 132 (31.8%) | 308 (22.5%) | 307 (27.0%) |
| Diabetes mellitus | 394 (24.8%) | 65 (15.9%) | 235 (20.3%) | 125 (30.1%) | 271 (19.8%) | 338 (29.8%) |
| CKD | 463 (29.2%) | 94 (23.0%) | 125 (10.8%) | 75 (18.1%) | 187 (13.7%) | 291 (25.6%) |
| Malignancy | 189 (11.9%) | 77 (18.8%) | 90 (7.8%) | 47 (11.3%) | 124 (9.1%) | 124 (10.9%) |
| **Critical illness and severity** |  |  |  |  |  |  |
| SOFA score (24 h) | 5.00 (3.00–8.00) | 6.00 (4.00–9.00) | 2.00 (1.00–4.00) | 4.00 (2.00–6.00) | 4.00 (2.00–6.00) | 4.00 (2.00–7.00) |
| OASIS score | 34.00 (28.0–40.0) | 33.00 (27.0–39.0) | 28.00 (23.0–34.0) | 30.00 (25.0–36.0) | 32.00 (26.0–37.0) | 33.00 (27.0–38.0) |
| ICU LOS, days | 4.17 (2.88–8.21) | 3.83 (2.75–6.96) | 3.50 (2.58–5.21) | 3.54 (2.54–5.25) | 3.58 (2.63–5.88) | 3.83 (2.67–6.76) |
| Hospital LOS, days | 13.00 (8.00–22.0) | 11.00 (7.00–20.0) | 7.00 (4.00–10.00) | 8.00 (6.00–13.50) | 8.00 (6.00–13.00) | 11.00 (7.00–19.0) |
| Sepsis (Sepsis-3) | 862 (54.3%) | 220 (53.8%) | 324 (28.0%) | 145 (34.9%) | 578 (42.3%) | 551 (48.5%) |
| **In-hospital therapies** |  |  |  |  |  |  |
| Surgery | 1,010 (63.6%) | 177 (43.3%) | 433 (37.4%) | 148 (35.7%) | 740 (54.2%) | 664 (58.5%) |
| PRBC transfusion | 659 (41.5%) | 124 (30.3%) | 45 (3.9%) | 60 (14.5%) | 241 (17.6%) | 338 (29.8%) |
| PRBC transfusion, units | 1.64 ± 4.03 | 1.00 ± 2.66 | 0.14 ± 0.83 | 0.34 ± 1.14 | 0.73 ± 2.68 | 0.97 ± 2.27 |
| Vasopressor use (first 48 h) | 685 (43.2%) | 155 (37.9%) | 176 (15.2%) | 105 (25.3%) | 394 (28.8%) | 441 (38.8%) |
| Invasive mechanical ventilation^&^ | 946 (66.3%) | 179 (53.3%) | 373 (44.0%) | 152 (46.1%) | 656 (56.9%) | 606 (60.4%) |
| **1-year mortality** | 368 (23.2%) | 154 (37.7%) | 137 (11.8%) | 93 (22.4%) | 212 (15.5%) | 249 (21.9%) |

Data are presented as median (interquartile range), mean ± standard deviation, or number (percentage), as appropriate.

^%^ Missing data: 43%; ^&^ Missing data: 16%

**Abbreviations:** BMI, body mass index; CKD, chronic kidney disease; COPD, chronic obstructive pulmonary disease; ICU, intensive care unit; MCH, mean corpuscular hemoglobin; MCHC, mean corpuscular hemoglobin concentration; MCV, mean corpuscular volume; OASIS, Oxford Acute Severity of Illness Score; PRBC, packed red blood cells; RDW, red cell distribution width; RBC, red blood cell; SOFA, Sequential Organ Failure Assessment

**eTable 3. Hematologic clusters derived from RBC indices at hospital discharge and association with one-year mortality**

| Risk category | Cluster^*^ | Erythropoietic characteristics^**^ | | Clinical characteristics^***^ | Adjusted HR (95% CI) ^****^ | | Hypothesis-generating erythroid profiles |
| --- | --- | --- | --- | --- | --- | --- | --- |
|  |  |  |  |  | **Derivation** | **Internal validation** |  |
| High | 1 | RDW↑↑,  MCV↑↑, Hb↓,  MCH↑, MCHC≈, RBC↓ | Macrocytic, high-RDW dysregulated erythropoiesis | High burden of malignancy and liver disease; higher illness severity | 2.14 (1.81–2.53) ^&^ | 2.28 (1.79–2.90) ^&^ | Overlap of chronic disease–associated ineffective erythropoiesis and acute illness–associated erythropoietic stress |
| Intermediate | 3 | RDW↑↑,  MCV↓↓, Hb↓, MCH↓↓, MCHC↓, RBC≈ | Microcytic / hypochromic high-RDW erythropoiesis | Older age;  chronic cardiometabolic disease | 1.87 (1.56–2.25) ^&^ | 1.56 (1.20–2.04) ^&^ | Iron-restricted or inflammatory chronic ineffective erythropoiesis |
|  | 5 | RDW↑,  MCV≈, Hb↓,  MCH≈, MCHC≈, RBC↓ | Normocytic anemia with elevated RDW | Sepsis common; moderate ICU instability and transfusion exposure | 1.68 (1.45–1.95) ^&^ | 1.46 (1.17–1.81) ^&^ | Acute illness–associated erythropoietic stress |
|  | 0 | RDW↑,  MCV≈/↑, Hb↓↓,  MCH≈, MCHC≈, RBC↓↓ | Severe normocytic anemia with high RDW | High comorbidity burden;  highest transfusion exposure | 1.57 (1.35–1.81) ^&^ | 1.42 (1.15–1.76) ^&^ | Anemia of chronic disease |
| Low | 4 | RDW≈,  MCV≈, Hb≈/↓,  MCH≈, MCHC≈, RBC≈↓ | Preserved / compensated erythropoiesis | More frequent surgical admissions; lower comorbidity burden | 1.21 (1.04–1.41) ^&^ | 1.10 (0.89–1.37) ^#^ | Preserved erythropoietic recovery at discharge |
|  | 2 | RDW≈,  MCV≈, Hb≈,  MCH≈, MCHC≈, RBC≈ | Preserved erythropoiesis | Lower comorbidity burden and critical illness severity | 1.00 (reference) | 1.00 (reference) | Preserved erythropoiesis at discharge |
| ^*^ Clusters were derived exclusively using pre-discharge red blood cell indices: RDW, hemoglobin, RBC count, MCV, MCH, and MCHC.  ^**^ Arrow notation indicates relative differences compared with the overall cohort: ↑ increased, ↓ decreased, ≈ approximately normal.  ^***^ Clinical characteristics were not used in cluster derivation and are shown for *post hoc* characterization only.  ^****^ Adjusted hazard ratios were estimated using Cox proportional hazards models with cluster 2 (lowest mortality risk) as the reference group. ^&^ p-value < 0.05 ^#^ p-value = 0.38  Abbreviations: CI, confidence interval; Hb, hemoglobin; HR, hazard ratio; MCH, mean corpuscular hemoglobin; MCHC, mean corpuscular hemoglobin concentration; MCV, mean corpuscular volume; RDW, red cell distribution width; RBC, red blood cell. | | | | | | | |

Qualitative assessment of RBC indices alongside clinical characteristics suggested distinct patient clusters that broadly aggregated into three strata of 1-year mortality risk: low, intermediate, and high. These clusters indicated erythroid profiles at hospital discharge that differed according to age, diagnosis at ICU admission (e.g., sepsis), acute illness severity, and comorbidities. Cluster 0 was characterized by notable anemia, modest RDW elevation, older age, and multiple comorbidities, a pattern potentially compatible with inflammation-associated chronic anemia. Cluster 1 showed anemia with higher RDW and markedly increased MCV in younger patients with more incident hepatic disease and cancer, possibly reflecting ineffective or inflammation-suppressed erythropoiesis. Cluster 2 demonstrated preserved hemoglobin levels with normal RDW in patients with low acute illness stress, potentially reflecting preserved marrow function and erythropoiesis. Cluster 3 exhibited moderate anemia with low MCV, MCHC, and MCH and markedly elevated RDW, a pattern that may reflect dysregulated erythropoiesis related to chronic disease and micronutrient deficiency, including iron-homeostasis disorders. Cluster 4 was characterized by higher hemoglobin and RBC mass among predominantly surgical patients with lower comorbidities, possibly compatible with preserved erythropoietic reserve despite acute traumatic stress. Cluster 5 showed modest RDW elevation, lower hemoglobin, frequent RBC transfusion exposure, and higher sepsis prevalence, potentially reflecting dynamic hematological instability during critical illness. Similar findings were observed in the validation cohort. Clinical interpretation of these clusters should remain exploratory, requiring further validation and mechanistic evaluation in dedicated studies.
